# The Trans-omics Landscape of COVID-19

**DOI:** 10.1101/2020.07.17.20155150

**Authors:** Peng Wu, Dongsheng Chen, Wencheng Ding, Ping Wu, Hongyan Hou, Yong Bai, Yuwen Zhou, Kezhen Li, Shunian Xiang, Panhong Liu, Jia Ju, Ensong Guo, Jia Liu, Bin Yang, Junpeng Fan, Liang He, Ziyong Sun, Ling Feng, Jian Wang, Tangchun Wu, Hao Wang, Jin Cheng, Hui Xing, Yifan Meng, Yongsheng Li, Yuanliang Zhang, Hongbo Luo, Gang Xie, Xianmei Lan, Ye Tao, Hao Yuan, Kang Huang, Wan Sun, Xiaobo Qian, Zhichao Li, Mingxi Huang, Peiwen Ding, Haoyu Wang, Jiaying Qiu, Feiyue Wang, Shiyou Wang, Jiacheng Zhu, Xiangning Ding, Chaochao Chai, Langchao Liang, Xiaoling Wang, Lihua Luo, Yuzhe Sun, Ying Yang, Zhenkun Zhuang, Tao Li, Lei Tian, Shaoqiao Zhang, Linnan Zhu, Lei Chen, Yiquan Wu, Xiaoyan Ma, Fang Chen, Yan Ren, Xun Xu, Siqi Liu, Jian Wang, Huanming Yang, Lin Wang, Chaoyang Sun, Ding Ma, Xin Jin, Gang Chen

## Abstract

System-wide molecular characteristics of COVID-19, especially in those patients without comorbidities, have not been fully investigated. We compared extensive molecular profiles of blood samples from 231 COVID-19 patients, ranging from asymptomatic to critically ill, importantly excluding those with any comorbidities. Amongst the major findings, asymptomatic patients were characterized by highly activated anti-virus interferon, T/natural killer (NK) cell activation, and transcriptional upregulation of inflammatory cytokine mRNAs. However, given very abundant RNA binding proteins (RBPs), these cytokine mRNAs could be effectively destabilized hence preserving normal cytokine levels. In contrast, in critically ill patients, cytokine storm due to RBPs inhibition and tryptophan metabolites accumulation contributed to T/NK cell dysfunction. A machine-learning model was constructed which accurately stratified the COVID-19 severities based on their multi-omics features. Overall, our analysis provides insights into COVID-19 pathogenesis and identifies targets for intervening in treatment.

## Introduction

Coronavirus disease 2019 (COVID-19), a newly emerged respiratory disease caused by severe acute respiratory syndrome coronavirus 2 (SARS-CoV-2), has recently become a pandemic (WHO, 2020). COVID-19 is now found in almost all countries, totaling 11 327 790 confirmed cases and 532 340 deaths worldwide as of July 6th, 2020 (Worldometers, 2020).

The symptoms of COVID-19 vary dramatically ranging from asymptomatic to critical. Several studies have reported on confirmed patients who exhibit no symptoms (i.e., asymptomatic) (Bai et al., 2020; Chan et al., 2020; Lu et al., 2020b; Pan et al., 2020). Since such individuals are not routinely tested, the proportion of asymptomatic patients is not precisely known, but appears to range from 13% in children (Dong et al., 2020) to 50% in the testing of contact tracing evaluation(Kimball et al., 2020). Of COVID-19 patients with symptoms, 80% are mild to moderate, 13.8% are severe, and 6.2% are classified as critical (WHO, 2020; Wu and McGoogan, 2020). Furthermore, although the overall mortality rate of diagnosed cases was estimated to be ∼3.4% (Worldometers, 2020), the rate varied from 0.2% to 22.7% depending on the age group and other health issue (Novel Coronavirus Pneumonia Emergency Response Epidemiology, 2020; Onder et al., 2020). Some confounding factors appear to be associated with COVID-19 progress and prognosis. For example, preliminary evidence suggests that comorbidities such as, hypertension, diabetes, cardiovascular disease, and respiratory disease results in a worse prognosis of COVID-19 (Zheng et al., 2020), and dramatically increase mortality (Novel Coronavirus Pneumonia Emergency Response Epidemiology, 2020). Therefore, the pathogenesis and mortality caused by SARS-CoV-2 infection in otherwise healthy individuals are not clear. Death due to COVID-19 is significantly more likely in older patients (i.e.,≥65 years old), possibly due to the decline in immune response with age (Wu et al., 2020a; Zheng et al., 2020).

So far, most studies have focused on the relationship between the disease and clinical characteristics, sequencing of virus genomes (Lu et al., 2020a) and identifying the structure of the SARS-CoV-2 spike glycoprotein (Lan et al., 2020; Walls et al., 2020). There has been some work on integrated multi-omics signatures. For example, meta-transcriptome sequencing was conducted on the bronchoalveolar lavage fluid of SARS-CoV-2 infected patients (Xiong et al., 2020). Proteomic and metabolomic analyses of the serum from COVID-19 patients have also been investigated (Bojkova et al., 2020; Shen et al., 2020; Wu et al., 2020b). However, systematic study of COVID-19 remains lacking. From data so far, it is difficult to determine which parameters are due to the infection and which to the comorbidities.

Here, to focus on the sole effect of SARS-CoV-2 infection on disease severity, 231 COVID-19 cases with different clinical severity and without comorbidities were selected. We performed trans-omics analysis, which included genomic, transcriptomic, proteomic, metabolomic, and lipidomic analytes of blood samples from COVID-19 patients, to better understand the associations among genetics and molecular mechanisms of consecutively severe COVID-19. We proposed a novel mechanism for inflammatory cytokine regulation at the post-transcriptional level. Cytokine storm, tryptophan metabolites, and T/NK cell dysfunction cooperatively contribute to the severity of COVID-19.

## Results

### Patient Enrollment and Trans-omics Profiling for COVID-19

To gain a comprehensive insight into the molecular characteristics of COVID-19 in patients characterized with different disease severity, a cohort of 231 out of 1432 COVID-19 patients were selected based on stringent criteria for the trans-omics study (**Figure S1**). Given that older age and comorbidities appear to have effects on disease progression and prognosis (Guan et al., 2020; Zhang et al., 2020; Zhou et al., 2020), participants aged between 20 and 70 years old (mean±SD, 46.7±13.5) without comorbidities were selected, to minimize the impact of confounding factors. Detailed information about the enrolled patients, including sampling date and basic clinical information, was shown in **Figure S2,** and **Table S1** and **S2**, Among the enrolled 231 COVID-19 patients, 64 were asymptomatic, 90 were mild, 55 were severe, and 22 were critical. In-depth multi-omics profiling was performed, including whole-genome sequencing (203 samples) and transcriptome sequencing (RNA-seq and miRNA-seq of 178 samples) of whole blood. Concurrently, liquid chromatography–mass spectrometry (LC-MS) was performed to capture the proteomic, metabolomic, and lipidomic features of COVID-19 patient sera (161 samples) (**Figure 1A**). After data pre-processing and annotation, the final dataset contained a total of 25882 analytes including 18245 mRNAs, 240 miRNAs, 5207 lncRNAs, 634 proteins, 814 metabolites, and 742 complex lipids (**Figure 1B, Figure S3** and **Table S3.1**). To quantify the molecular profiles in relation to disease severity, we conducted pairwise comparisons between the four severity groups for each omics-level (see **Methods**). Results indicated extensive changes across all omics levels (**Figure 1B, Figure S4** and **Table S3.2-3.5**). The percentage of analytes that changed dramatically in at least two comparisons ranged from 24.18% (mRNAs) to 54.57% (proteins) (**Figure 1B**). Generally, we first found profound differences between asymptomatic and symptomatic patients at all omics levels, suggesting a specific molecular feature in this particular population. Second, the changes in analytes between mild and severe were subtle at all omics levels except for protein, indicating marked molecular similarities between these two severities, even in the presence of differences in clinical manifestations. Third, the differences between the critical group and other groups were extremely high, implying a sudden and dramatic change from severe to critical disease.

**Figure 1.**
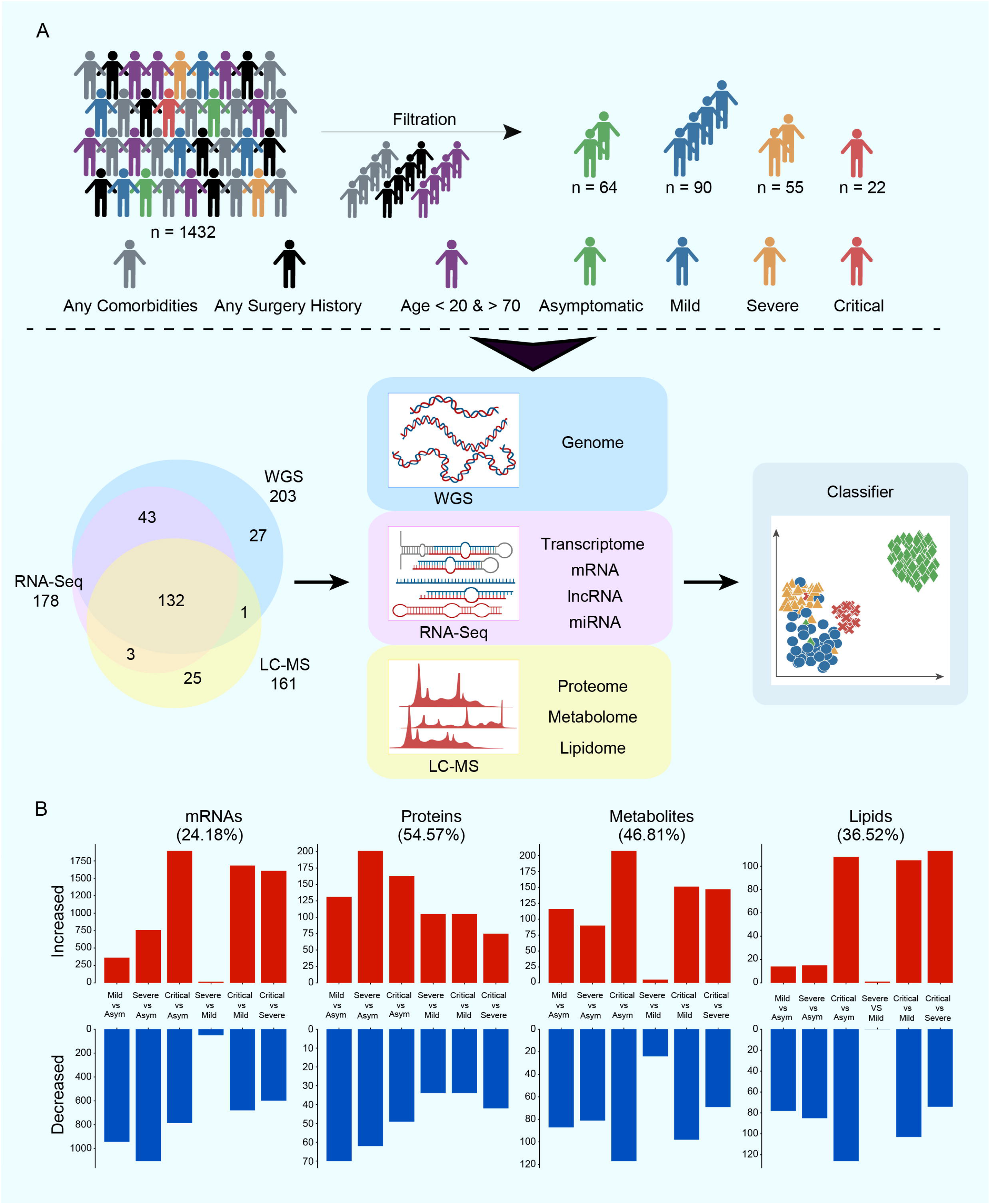
Patient Enrollment, Study Design and Trans-omics Profile of COVID-19 Severity. (A) Overview of patient enrollment criteria and the study design including multi-omics profiling from blood samples of COVID-19 patients spanning four disease severities including Asym (short for asymptomatic), Mild, Severe and Critical. Venn diagram showing the overlapping of final samples pass QC for each high throughput method. Classifier showing machine learning prediction model construction (B) Multi-omics changes among each disease severity group. See also **Figures S1-S2** and **Tables S1-S2.**

### Genomic Architecture of COVID-19 Patients

Based on whole-genome sequencing of 203 unrelated patients, we obtained a total of 18.9 million high quality variants, in which 15.3 million bi-allelic single nucleotide polymorphisms (SNPs) were used in the following analyses (**Figures S5A**-**H** and **Table S4.1**). Principal component analysis showed no obvious population stratification within the study and the patients were grouped together with East Asian and Han Chinese when compared to 1000GP (Auton et al., 2015) phase 3 released data (**Figures S5I**-**J**). Single-variant based association tests were performed to investigate the connections among common variants (MAF>0.05) and the diversity of clinical manifestations. We first compared the generalized severe group (severe and critical, n=65) with the mild group (asymptomatic and mild, n=138) (**Figures S6A**-**B**), then compared the asymptomatic group (n=63) with all other symptomatic patients (n=140) (**Figures S6C**-**D, Table S4.2**). Gender, age, and top 10 principal components were included as covariates. In general, no signal showed genome-wide significance (*P* <5e^-8^) in these comparisons. A suggestive signal (*P* <1e^-6^) associated with the absence of symptoms was found on chromosome 20q13.13, which comprised six SNPs, the most significant being SNP rs235001 **(Table S4.3)**. Locus zoom identified two protein coding genes *B4GALT5* and *PTGIS* in the region spanning ±50k of the SNP (**Figure S6E**). Considering the small sample size of this study, the associated SNPs still need to be confirmed by further investigations. We also assessed two loci, rs657152 at locus 9q34.2 and rs11385942 at locus 3p21.31, which have been found to be associated with severe COVID-19 with respiratory failure in Spanish and Italian populations (Ellinghaus et al., 2020). For rs657152, the overall frequency of the protective allele C was 0.5468 (222/406) in our data, which decreased in the critical group (AF=0.382, 13/34, Fisher’s exact test *P*=0.04896). For rs11385942, the risk allele GA was not detected in any patient in our study, as this variant was rare in Chinese people (Liu et al., 2018) (**Table S4.4**), consistent with previously reported global distribution (Ellinghaus et al., 2020).

Quantitative trait locus (QTL) analysis has been widely applied to infer the contribution of genetic variations to complex phenotypes (Fagny et al., 2017). Here, QTL analysis was performed to explore the correlations of proteomic, metabolomic and lipidomic features with genetic variations, resulting in 1328 mRNAs, 76 proteins, 195 metabolites and 4 lipids significantly associated with a variety of QTL (*P*≤5e^-8^) (**Table S5**). Taken together, we revealed the overall contribution of genetic variations to the output at different omics levels in COVID-19 patients.

### Changing Patterns of Transcriptome in Relation to COVID-19 Severity

Overall, 1302, 1862 and 2678 differentially expressed genes (DEGs) were identified in mild, severe and critical groups, compared to the asymptomatic group, among which 578 DGEs were shared by symptomatic groups. Within symptomatic groups, only 66 DEGs between severe and mild groups were identified while over 2000 DEGs were detected in critical compared to severe or mild, indicating a similar molecular feature between mild and severe and extremely distinct features between critical and the mild/severe groups at transcriptome level (**Figure 2A, Table S3.2**). To characterize progressive changes through the four disease severities of COVID-19, we conducted unsupervised clustering of mRNAs that were differentially expressed in at least three of the six comparison groups. At the mRNA level, we classified genes into three clusters according to their expression patterns across different disease severities (**Figure 2B, Table S6.1**). Intriguingly, the expression levels of genes in cluster 1 were increased both in asymptomatic and critically ill patients as compared to mild/severe patients, while the extend of upregulation was more profound in asymptomatic cases. GO analysis showed these genes were related to neutrophil activation, inflammatory response, granulocyte chemotaxis, and IL2, IL-6, IL-8 production (**Figure 2B, Table S6.2**). Key chemokines (CXCL8, CXCR1, CXCR2) for neutrophil activation and accumulation, as well as inflammatory responses genes (TLR4 and TLR6) associated with toll-like receptors, and several key inflammatory response genes (MMP8, MMP9, S100A12, S100A8, UBE2E3) shared this expression pattern (**Figure 2C**), suggesting a highly activated innate immune and pro-inflammatory response both in asymptomatic and critically ill patients than that in mild and severe patients at transcriptomic level.

**Figure 2.**
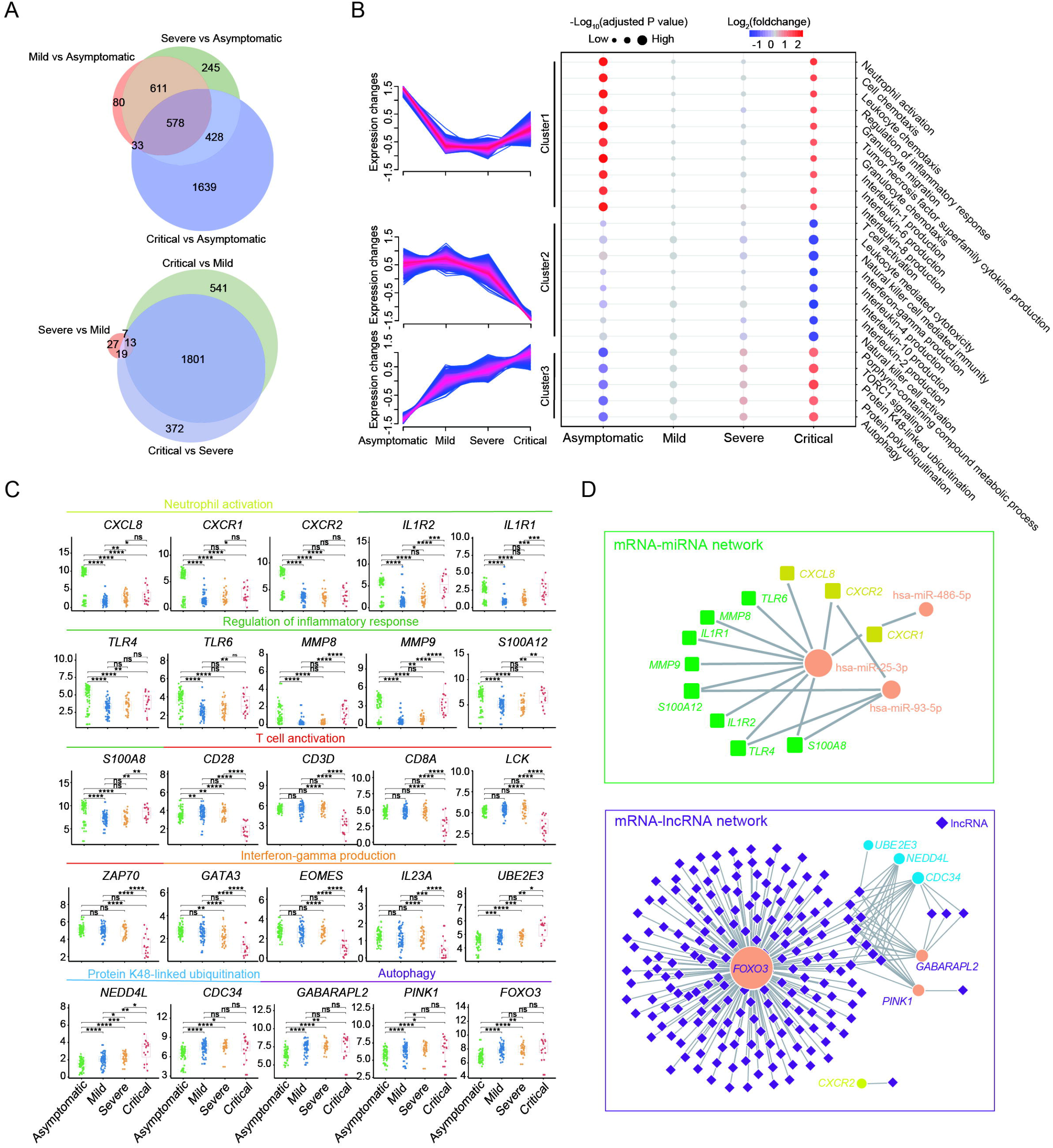
Gene Expression Changes through Disease Severity. (A) Venn diagram showing the overlapping of genes that are significantly altered in three symptomatic groups (mild, severe and critical) compared to asymptomatic group, and genes that are altered between each pairwise comparison of the three symptomatic groups. (B) GO enrichment analysis using genes in each cluster showing different changing patterns through progressive disease severity. GO direction is the median log2 fold change relative to mild of significant genes in each GO term (blue, downregulated; red, upregulated). The dot size represents GO significance. (C) Gene expression changes across four severity groups showing dynamic genes in T cell activation, interferon-gamma production, regulation of inflammatory response, regulation of inflammatory response and protein K48-linked ubiquitination. (D) miRNA-mRNA and lncRNA-mRNA interaction networks showed regulations of miRNAs and lncRNAs of the dynamic genes. See also **Figures S1-S2** and **Tables S3.2, S6.1-S6.2, S7-S8.**

Genes in cluster 2 were enriched in T cell activation, leukocyte-mediated cytotoxicity, NK cell-mediated immunity, and interferon-gamma production. The expression levels of these genes were specifically decreased in critical patients compared to the other three severities. Important genes for T cell activation, such as CD28, LCK, and ZAP70, as well as key transcript factors for interferon-gamma production (GATA3, EOMES and IL23A), showed this expression pattern across the different disease severities (**Figure 2C**). Thus, although innate immune was activated in both asymptomatic and critically ill patients, T cell mediated adaptive immune response was specifically suppressed in critical COVID-19 patients.

Cluster 3 contained genes primarily involved in protein polyubiquitination and autophagy. The expression of genes in this cluster gradually increased from the asymptomatic to mild/severe and then peaked at the critical (**Figure 2B**). An important transcript factor for autophagy, FOXO3, showed this expression pattern (**Figure 2C**). Genes in this cluster reflected the increasing tissue damage and cell death along with disease severity.

### Post-transcriptional Regulation by Non-coding RNAs in Relation to COVID-19 Severity

Next, we investigated the post-transcriptional regulatory network associated with the genes in **Figure 2C**. From 240 high abundant miRNAs, 625 pairs of mRNA-miRNA were selected, of which 16 pairs were with coefficients < −0.5 (**Table S7**). The expression of three miRNAs (miR-25-3p, miR-486-5p and miR-93-5p) was uncovered to be negatively correlated with that of eleven genes including eight inflammatory response genes, and three neutrophil activation genes (**Figure 2D**). Meanwhile, 5207 lncRNAs were identified from the NCBI database, and 3084 pairs of mRNA-lncRNA connection were tested (**Table S8**). After removing low correlation coefficients, 233 pairs of mRNA-lncRNA connection with coefficients ≤-0.6 were left, and we found many lncRNAs to be strongly and negatively correlated with FOXO3 which plays a critical role in autophagy (**Figure 2D**) (Mammucari et al., 2007). In addition, two autophagic genes, PINK1 and GABARAPL2, were found to be correlated with expression level of lncRNA as well. In view of the fact that the expression of FOXO3, a negative regulator of the antiviral response, elevated along with the aggravation of the patient’s condition, lncRNA differential accumulation might play a role in autophagy and antiviral response dysregulation in critically ill COVID-19 patients (**Figure 2C**) (Litvak et al., 2012).

### Changing Patterns of Proteins, Metabolites and Lipids in Relation to COVID-19 Severity

Generally, all proteins, metabolites and lipids were classified into seven clusters with four progressive severities: increasing patterns, decreasing patterns, U-shaped patterns and stage specific patterns. Increasing patterns include the gradually increasing cluster C2 and the sharply increasing cluster C3. Decreasing patterns were composed of gradually decreasing cluster C6 and sharply decreasing cluster C1. Additionally, C4, C5 and C7 belonged to the U-shaped patterns, mild specific and critical specific patterns respectively (**Figure 3A, Figure S7** and **Table S9.1**). To systematically characterize the interaction networks among proteins, metabolites and lipids within each cluster, we conducted co-expression network analysis using ranked spearman correlation coefficient (see **Methods**), resulting in a systematic multi-omics network for each cluster (**Figure S8, Tables S10.1**-**10.2)**. Overall, we revealed putative dynamic interactions within each network, connecting immunity proteins (CSF1, C1S *etc.*) to specific groups of metabolites (phenylalanine, tryptophan *etc.*) and lipids (phosphatidylethanolamine, triglyceride *etc.*).

**Figure 3.**
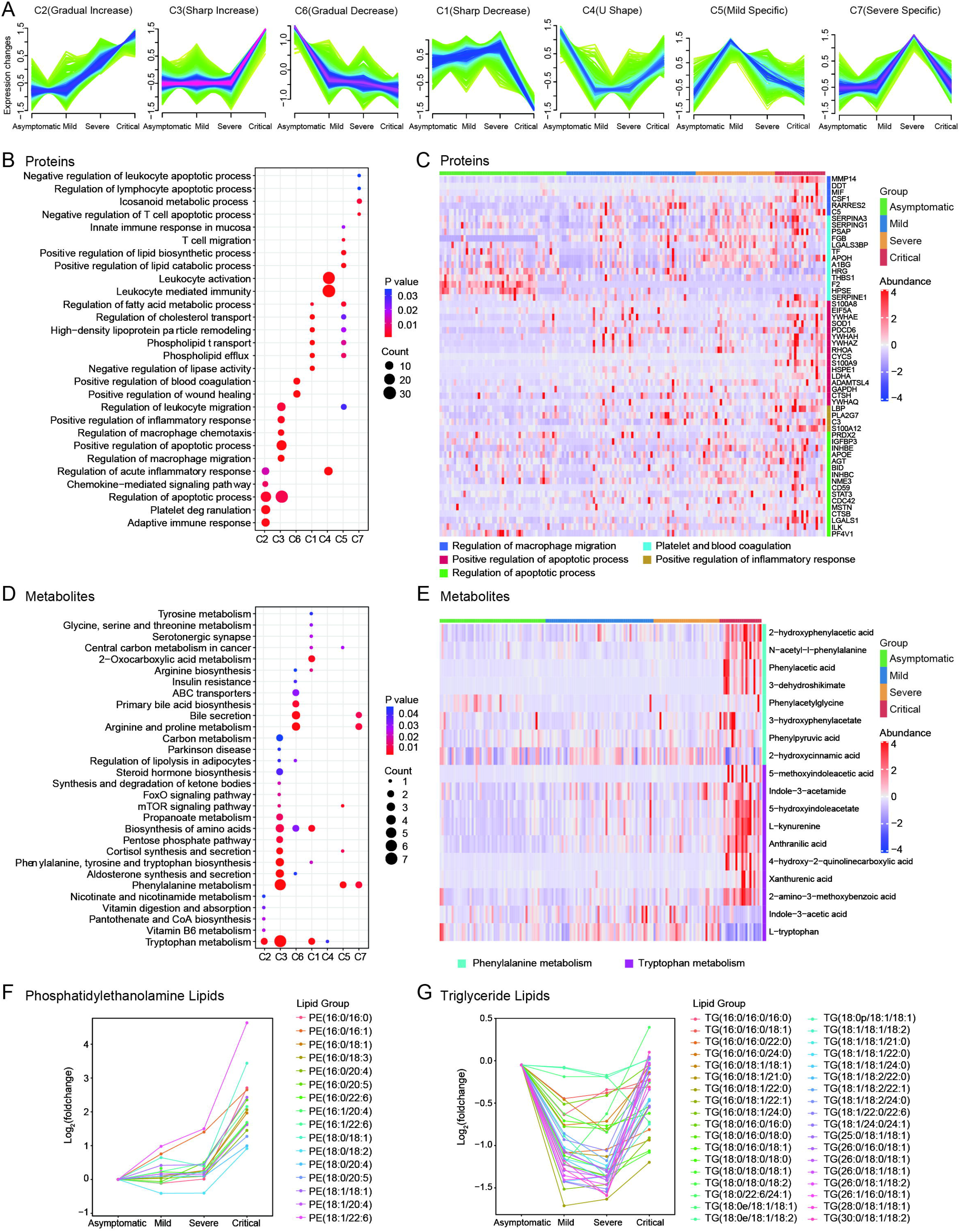
Multi-omic Changes in Proteins, Metabolites and Lipids across Four Severity Groups. (A) Clustering of longitudinal trajectories using circulating plasma analytes including proteins, metabolites and lipids (FDR <0.05). (B) Enriched GO Biological Process (BP) terms for proteins in seven clusters. (C) Heatmap of representing protein expression in three functional categories. Each column indicates a patient sample and row representes proteins. Color of each cell shows Z-score of log2 protein abundance in that sample. (D) Enriched KEGG pathway for metabolites in seven clusters. (E) Heatmap of representing metabolites expression in Phenylalanine and Tryptophan metabolism. (F) Dynamic expression changes in Phosphatidylethanolamine lipids across four disease severity groups. The dots represent the log2 fold change relative to asymptomatic for each lipid in the group. (G) Dynamic expression changes in Triglyceride lipids across four disease severity groups. See also **Figures S7-S9** and **Table S2, S9.1-S9.3** and **S10.1-10.2.**

Notably, a variety of biological pathways found to be specifically enriched in the different clusters **(Figure 3B, Table S9.2)**. We were particularly interested in proteins showing high expression levels in the two extreme severities (i.e., asymptomatic, and critical patients). Consistent with transcription analysis (**Figure 2B**), a variety of proteins (BID, ILK, ADAMTSL4 *etc*.) related to the positive regulation of apoptotic processes were preferentially present in critical COVID-19 patients. However, inconsistent with mRNA expression patterns, proteins associated with positive regulation of inflammatory response and macrophage migration (S100A8, S100A12, C5, LBP, DDT *etc.*) were only activated in critically ill patients but not asymptomatic patients (**Figure 3C**). Proteins associated with platelet and blood coagulation (F2, HPSE, PF4 *etc*.) showed a decreasing pattern from asymptomatic to severe patients. (**Figure 3C**), supporting the observed thrombocytopenia and coagulopathy in critically ill patients.

Metabolites also showed distinct profiles in the different clusters. In particular, compared to other severities, increase in phenylalanine and tryptophan biosynthesis was observed in critical COVID-19 patients (**Figures 3D**-**E, Table S9.3**). Tryptophan metabolism was considered a biomarker and therapeutic target of inflammation (Sorgdrager et al., 2019) and changes in tryptophan metabolism were reported to be correlated with serum interleukin-6 (IL-6) levels (Moffett and Namboodiri, 2003). Consistently, we detected enrichment of IL-6 in critical patients using enzyme-linked immunosorbent assay (ELISA) (**Figure 3E, Table S2**).

We also investigated the dynamics of lipids among the different severities. Alterations in several major lipid groups and bioactive molecules were revealed in the symptomatic patients, especially in the critical group. Phosphatidylethanolamine (PE) including dimethyl-phosphatidylethanolamine (dMePE), sphingomyelin (SM), Lysophosphatidylinositol (LPI), Monoglyceride (MG), Sphingomyelin phytosphingosine (phSM), Phosphatidylserine (PS) were elevated (**Figure S9**). Notably, we found two groups of lipids showing specific expression patterns, the phosphatidylethanolamine lipids belonging to the gradually increasing cluster (cluster 1), and triglyceride lipids presented in the U-shaped cluster with elevated levels in the asymptomatic and critical patients. Consistent with previous findings that RNA virus replication is dependent on the enrichment of phosphatidylethanolamine distributed at the replication sites of subcellular membranes (Xu and Nagy, 2015), we observed that the expression patterns for a cohort of 16 phosphatidylethanolamine lipids positively correlated with COVID-19 severities (**Figure 3F**). Furthermore, we identified that a repertoire of 36 triglyceride lipids were relatively low in both mild and severe COVID-19 patients (**Figure 3G**). In addition to the above abundant structural lipid classes, several bioactive lipids also changed significantly in the symptomatic groups, including lysophosphatidylcholine (inhibiting endotoxin-induced release of late proinflammatory cytokine) (Yan et al., 2004) and lysophosphatidyliositol (an endogenous agonist for GPR55 whose activation regulates several pro-inflammatory cytokines) (Marichal-Cancino et al., 2017), suggesting that lipidome changes that interfere with cell membrane integrity and normal functions or disturb inflammatory and immune states may play important and complex roles in COVID-19 disease development (**Figure S9**).

### Prediction of COVID-19 Severity Using Machine Learning Model

Based on multi-omics analysis, we found that the mild and severe groups shared many similar characteristics. However, it is important to predict these two groups, which is important for early intervention and then preventing disease progress. We therefore developed an XGBoost (extreme gradient boosting) machine learning model by leveraging multi-omics data. We randomly stratified samples for the training set (80%) and the independent testing set (20%) (**Figure 4A**, see **Methods**). After normalization, a total of 297 multi-omics features were preliminarily selected by applying a hybrid method (see **Methods**). The XGBoost model trained based on these selected features achieved a mean micro-average AUROC (area under the receiver operating characteristic curve) and mean micro-average AUPR (area under the precision-recall curve) of 0.9715 (95% CI, 0.9497–0.9932) and 0.9495 (95% CI, 0.9086–0.9904) in the training set, respectively (**Figures 4B-C**). This showed strong generalizable discrimination among four severities based on 5-fold cross validation over 100 iterations.

**Figure 4.**
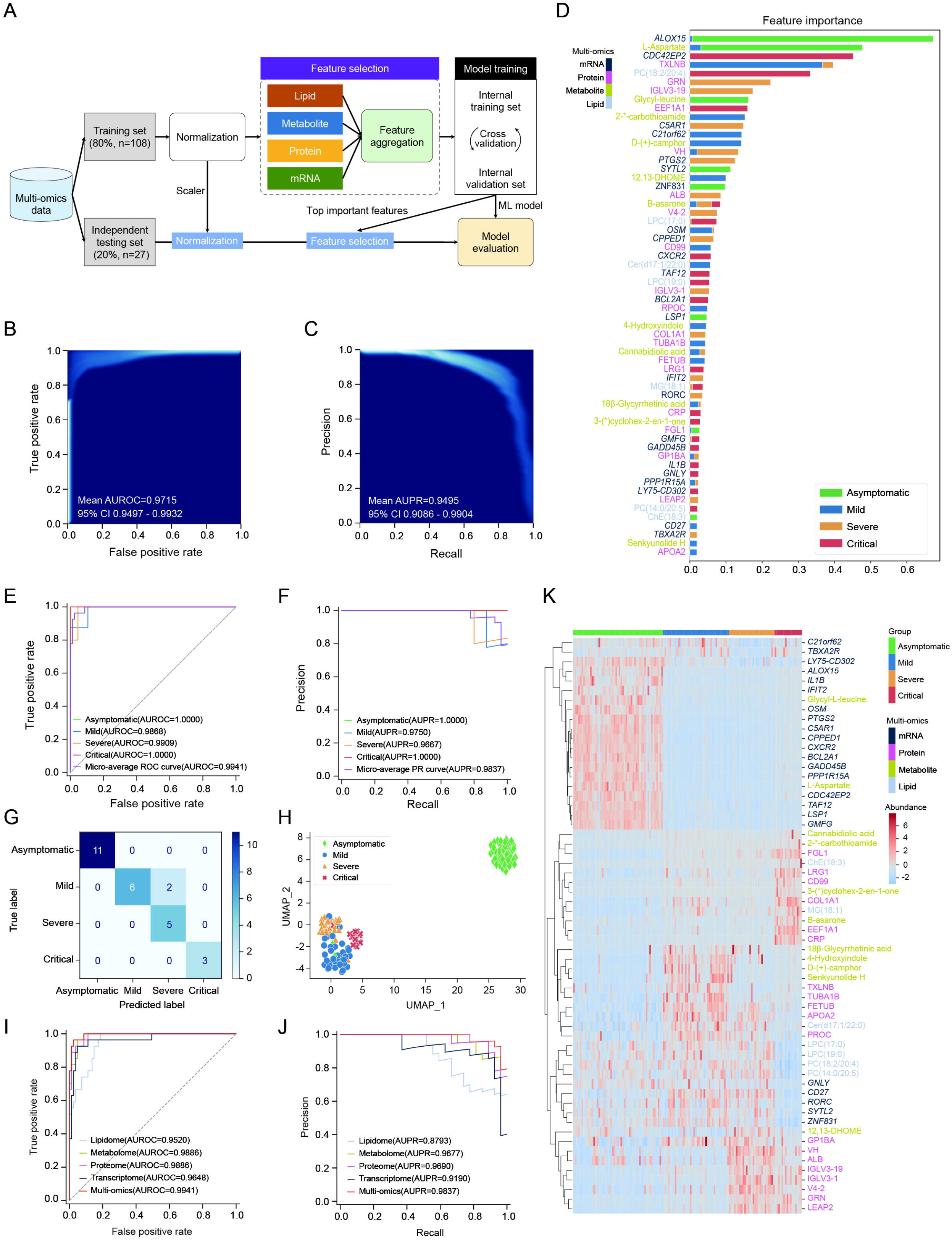
Performance of Machine Learning Model to Predict COVID-19 Patient Severity of Asymptomatic, Mild, Severe and Critical using Multi-omics Data. (A) Flowchart of developing XGBoost machine learning model. The model was trained with cross validation using training set (n=108) after normalization and feature selection, and re-trained with the identified top 60 important features. The re-trained model was further applied to assess generalization and performance using independent testing set (n=27). (B-C) Performance of the model learned in training set in terms of mean micro-average AUROC (B) and mean micro-average AUPR (C). Rasterized density plot of ROC (B) and PR (C) curve data from 5-fold cross validation for 100 iterations. (D) Top 60 important features (mRNA, n=23; protein, n=19; metabolite, n=11; lipid, n=7) ranked by SHAP value. The stacked bar indicated the average impact of the feature on the model output magnitude for different classes. (E-F) Performance of XGBoost model based on the top 60 features for distinguishing the 4 groups of COVID-19 severity in independent testing set in terms of AUROC (E) and A UPR (F). (G) Confusion matrix for predicting COVID-19 severity in independent testing set(n=27). (H) UMAP plot based on the top 60 features showing the distinct separation among the 4 types of COVID-19 severity in the whole data set (patients n=135). (I-J) Comparison of performance of models learned by each single-omics data with that of multi-omics data in independent testing set in terms of AUROC (I) and AUPR (J). (K) Heatmap of demonstrating the top 60 features profiles of the 4 groups of COVID-19 severity in the whole dataset (patients n=135). 2-*-carbothioamide: 2-(1-adamantylcarbonyl)hydrazine-1-carbothioamide, 3-(*)cyclohex-2-en-1-one: 3-(benzylamino)-5-(4-chlorophenyl)cyclohex-2-en-1-one See also **Figures S10∼S14**

The multi-omics features were prioritized and ranked by the XGBoost model and the SHAP (SHapley Additive exPlanations, see **Methods**) value and top 60 important features were selected, composed of 19 proteins, 11 metabolites, 7 lipids, and 23 mRNAs (**Figures 4D, S10-S13**). With the top 60 important features, XGBoost model was re-trained and validated, resulting in a micro-average AUROC and micro-average AUPR of 0.9941 and 0.9837 in the independent testing set, respectively (**Figures 4E**-**F**). The confusion matrix (**Figure 4G**) showed that all patients in the independent testing set were correctly identified, except for two mild patients who were predicted as severe. For further validation, we trained different XGBoost models through the same training protocol with each single-omics data. Results demonstrated that the XGBoost model outperformed that trained using single-omics features (**Figures 4I**-**J, S14**). Furthermore, we trained an additional XGBoost model based on the 24 features identified in Guo’s method (Shen et al., 2020) (two proteins and three metabolites were not detected in our experiment), leading to micro-average AUROC and micro-average AUPR in independent testing set are 0.9305 and 0.8300, respectively (**Figure S14**), which may be partially due to the different purposes for model construction, whereby Guo sought to distinguish severe patients from non-severe patients, whereas we attempted to identify four groups of COVID-19 patient severity. The UMAP (uniform manifold approximation and projection) plot showed distinct separation of disease severity groups, namely, asymptomatic, mild, severe, and critical (**Figure 4H**). Together, our results implied that the XGBoost model based on the top 60 multi-omics features could precisely differentiate COVID-19 patient severity status.

Notably, two transcription factor encoding genes (ZNF831 and RORC) closely associated with immune response (da Silveira et al., 2017; He et al., 2018) were identified by our model as important discriminative features. Besides, inflammatory response molecular (ALOX15, C5AR1 *etc.*), cytokine-mediated signaling pathway components (PTGS2, OSM *etc*.), leukocyte activation genes (CPPED1, GMFG *etc.*), apoptotic genes (BCL2A1, IFIT2, GADD45B *etc.*), a variety of anti-inflammatory factors (such as lipids of phosphatidylcholine, lysophosphatidylcholine *etc*.) were included in the selected 60 discriminative features. Moreover, most mRNAs were expressed highly in asymptomatic patients, lipids such as phosphatidylcholine and lysophosphatidylcholine decreased considerably in critical group. Most proteins among the 60 features such as C-reactive protein (CRP) and EEF1A1 were expressed highly in critical patients (**Figure 4K**). These results demonstrated that our model could not only be employed to stratify COVID-19 patients, but also discover molecular associated with pathogenesis of COVID-19.

## Discussion

With the global outbreak of SARS-CoV-2, COVID-19 has become a serious, worldwide and public health concern. However, comprehensive analysis of multi-omics data within a large cohort remains lacking, especially for patients with various severity grades, i.e., asymptomatic across the course of the disease to critically ill. To the best of our knowledge, this is the first trial designed to systematically analyze trans-omics data of COVID-19 patients with grade of clinical severity. Furthermore, it is worth emphasizing that we excluded all patients of extreme age or with comorbidities, to minimize bias due to confounding factors related to severity.

Asymptomatic patients have drawn great attention as these silent spreaders are hard to identify and cause difficulties in epidemic control (Long et al., 2020). Here, we demonstrated an unexpected transcriptional activation of the pro-inflammatory pathway and inflammatory cytokines (**Figure 2B** and **5A**). However, consistent with a recent report (Long et al., 2020), secretion of inflammatory cytokines such as IL-6 and IL-8 was extremely low in sera from the asymptomatic population **(Table S2).** In contrast, critically ill patients were characterized with excessive inflammatory cytokine production (**Figure 3C, Table S2**), whereas their transcription levels were only modestly elevated (**Figures 2B, 5A)**.

Typically, inflammatory cytokine production is tightly regulated both transcriptionally and post-transcriptionally (Mino and Takeuchi, 2018; Tanaka et al., 2014). Post-transcription of inflammation-related mRNAs is mainly regulated by RNA-binding proteins (RBPs), including ARE/poly-(U) binding degradation factor 1 (AUF1, HNRNPD), tristetraprolin (TTP, ZFP36), Regnase-1 (ZC3H12A), ILF3, ZNF692, ZCCHC11, FXR1, ELAVL1, and BRF1/2 (Carpenter et al., 2014). Interestingly, these RBPs involved in the degradation and destabilization of inflammatory cytokines were highly expressed in asymptomatic patients but showed extremely low expression in critical patients (**Figure 5B**). By recognizing inflammatory cytokine mRNA with stem-loop structures, RBPs can degrade or decay inflammatory cytokine mRNA. The balance of these actions elegantly controls inflammation intensity (Carpenter et al., 2014). AUF1 attenuates inflammation by destabilizing mRNAs encoding inflammatory cytokines, including IL-2, IL-6, TNF and IL-1β (Cathcart et al., 2013; Sadri and Schneider, 2009). As an anti-inflammatory protein, TTP destabilizes inflammatory mRNAs, such as GM-CSF, IL-2, and IL-6 (Taylor et al., 1996). Regnase-1 has a wide antiviral spectrum and efficiently inhibits the influenza A virus. Furthermore, Regnase-1 restrains inflammation by negatively regulating IL6 and IL17 mRNA stabilization (Garg et al., 2015; Omiya et al., 2020). Thus, Regnase-1 depletion facilitates severe systemic inflammation and virus replication. Accordingly, we propose that the observed discrepancy between cytokine mRNA and protein levels could be attributed to post-transcriptional mRNA stabilization mediated by RBPs (**Figure 5C**). Our data suggests a novel mechanism for inflammatory cytokine regulation at the post-transcriptional level, which explains the molecular mechanism of various clinical symptoms and suggests that RBPs could be a potential therapeutic target in COVID-19. However, additional functional researches will be required to ascertain their contribution toward the development of COVID-19.

**Figure 5.**
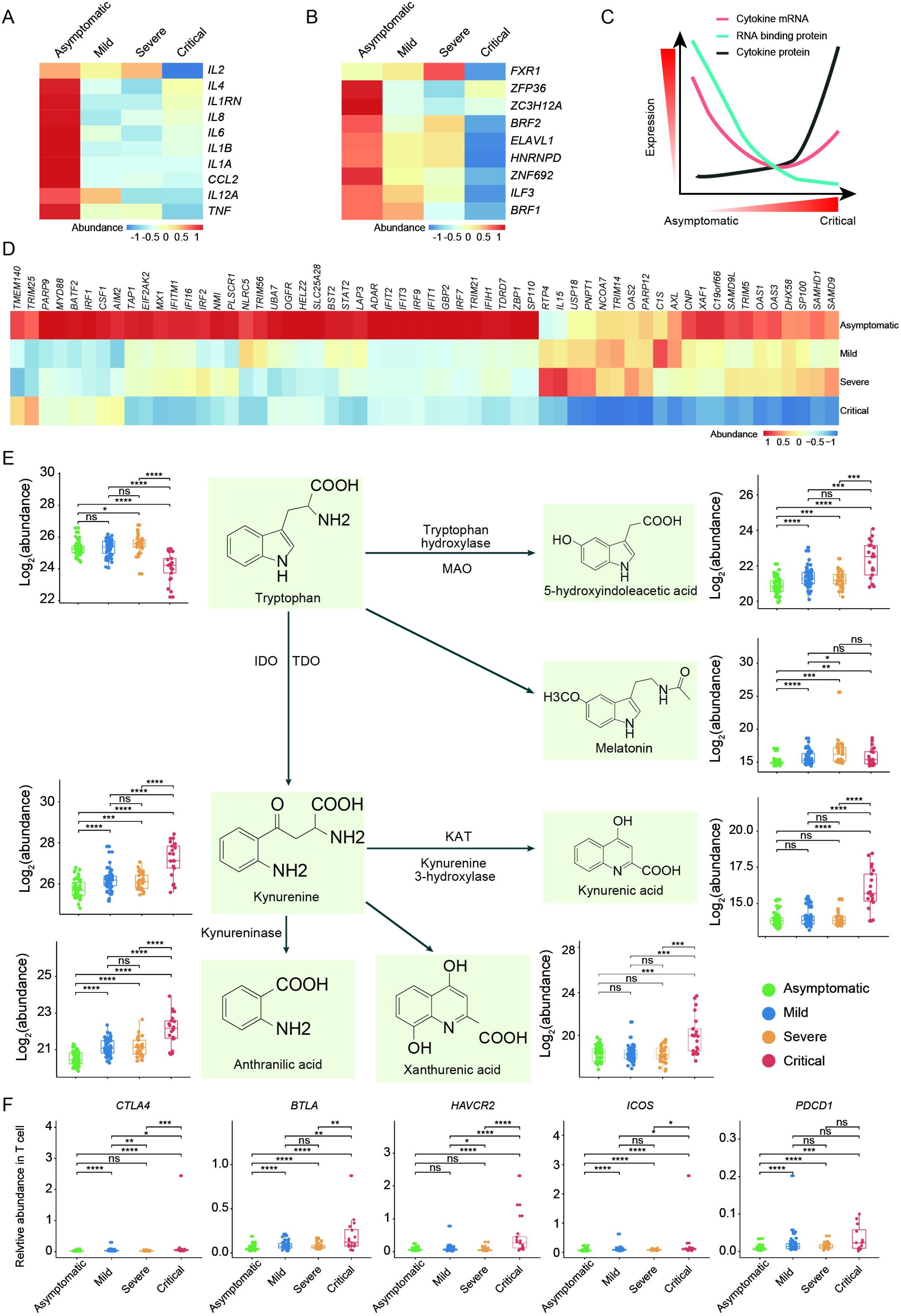
The novel insight of COVID-19 through Progressive Disease Severity. (A) Heatmap of demonstrating cytokines and chemokines DEGs expression across four disease severity. (B) Heatmap of demonstrating the expression levels of DEGs involved in RNA-binding proteins (RBP) across four disease severity. (C) The variation patterns of gene expression of cytokines and RBP, and immunological parameters across four disease severity. (D) Heatmap of demonstrating the expression levels of DEGs involved in IFN-1 responses across four disease severity. (E) Relative expression abundance of exhaustion marker genes *CTLA4, BTLA, HAVCR2, ICOS* and *PDCD1* in T cells. The relative expression abundance of the exhaustion marker genes was defined as their expression level dividing the expression level of T cell marker gene *CD3E*. (F) Summary of the pathways of tryptophan metabolism. IDO, indoleamine 2,3-dioxygenase; KAT, kynurenine aminotransferase; MAO, monoamine oxidase; TDO, tryptophan 2,3-dioxygenase. Box plots in this panel showed the expression level change (log2-scaled original value) of selected regulated metabolites across four disease severity.

An effective interferon (IFN) response can eliminate viral infection including that of SARS-CoV-2 (Bost et al., 2020). Insufficient activation of IFN signaling may contribute to severe cases of COVID-19 (Blanco-Melo et al., 2020; Broggi et al., 2020). As such, we compared the pathways of anti-viral IFN responses in the different severities of COVID-19 patients. Intriguingly, we found that critically ill patients failed to launch a robust IFN response compared with the highly activated IFN response observed in asymptomatic patients (**Figure 5D**). The impaired IFN response could be responsible for the loss of viral replication control in critically ill patients (Diao et al., 2020). Moreover, highly accumulated PE lipids (**Figure 3F**), which are important for RNA virus replication (Xu and Nagy, 2015), further enhanced SARS-CoV-2 replication. Consequently, uncontrolled viral replication could result in the orchestration of a much stronger immune response in critically ill patients, characterized by cytokine storms and immunopathogenesis. Conversely, the sufficient IFN response in asymptomatic patients could help to defend against viral infections.

Another feature of critically ill patients was defects in the T/NK cell-mediated adaptive immune response (**Figure 2B**), and accelerated tryptophan (Trp) metabolism (**Figure 5E**). In addition to the dramatically decreased T cell markers expression in critically ill patients (**Figure 2C**), we noticed a significant upregulation in exhaustion markers: *e.g*., PD-1, CTLA4, TIM3, ICOS, and BTLA in T cells (**Figure 5F**). T cell depletion is in line with the clinically observed T cell lymphopenia, which was also negatively correlated with COVID-19 severity. T cells play a critical role in antiviral immunity against SARS-CoV-2 (Grifoni et al., 2020), but their functional state and contribution to COVID-19 severity remain largely unknown. Recent research has demonstrated that SARS-CoV-2 dramatically reduces T cells, and up-regulates exhaustion markers PD-1, and Tim-3, especially in critically ill patients (Diao et al., 2020). Mechanistically, uncontrolled cytokine release may prompt the depletion and exhaustion of T cells. Clinically, T-cell counts are negatively associated with serum IL-6, IL-10 and TNF-alpha concentrations (**Table S2**) (Zhou et al., 2020). Also, it is well known that accelerated Trp metabolism by rate-limiting enzymes, i.e., indoleamine 2,3-dioxygenases (IDO1 and IDO2), mediates T cell dysfunction (Cronin et al., 2018). Tryptophan degradation products, such as L-kynurenine (Kyn), have an immunosuppressive function by depleting T cells and increasing apoptosis of T-helper 1 lymphocytes and NK cells (Mullard, 2018; Munn et al., 2005). Thus, we proposed that T cells also become metabolically exhausted and dysfunctional in critical patients due to the accelerated tryptophan (Trp) metabolism (**Figure 5E**).

It is possible that biological crosstalk exists among the cytokine storm, Trp metabolism, and T cell dysfunction processes. First, considering the essential role of Trp metabolism in blocking expansion and proliferation of conventional CD4^+^ helper T cells and effector CD8^+^ T cells and in potentiating CD4^+^ regulatory T (Treg) cell function(Cronin et al., 2018), accumulated Trp catabolite production, KYN, 3-HAA and Quin, inhibits adaptive T cell immunity. Second, Trp directly stimulates immune checkpoint expression levels, such as CTLA4 and PD-1 (Opitz et al., 2020). Third, in addition to the direct effects on T cell dysfunction, proinflammatory cytokines, e.g., IL-1β, IFN-γ, and IL-6, can lead to a robust elevation in circulating Kyn levels by up regulation of IDO/TDO (Wang et al., 2017), which synergistically worsen T cell dysfunction. Fourth, adaptive T cell immunity plays an unexpected role in tempering the initial innate response (Kim et al., 2007), T cells defection in critically ill patients could in turn exacerbate an uncontrolled innate immune response.

Therapeutically, considering the essential effects of tryptophan, IDO, and T cell function on COVID-19 severity, bolstering the immune system by restoring exhausted T cells may be a promising strategy for disease treatment. Targeting Trp catabolism by indoximod, or targeting IDO1/TDO2 by navoximod (NLG919) (Ricciuti et al., 2019), BMS-986205 (Gunther et al., 2019), or PF-06840003 (Crosignani et al., 2017) could metabolically restore T cell function. Furthermore, immune checkpoint blockage with PD1/PD-L1 or CTLA4 antibody increased T cell numbers and restore T cell function (Waldman et al., 2020), may be a potential strategy for treatment of critically ill patients. It may therefore be worthwhile to test if immune-boosting strategies are effective in COVID-19 clinical trials.

In this study, mild and severe groups shared common multi-omics features, with the exception of protein expression. However, it is crucial to distinguish mild and severe COVID-19 in clinical practice. For severe patients, oxygen facilities should be applied in the early stages to prevent progression to critical illness, who carries a much higher risk of death. Based on clinical needs, we applied a machine-learning prediction model in this study. Many prediction models have been used to assist medical staff to predict disease progression and outcome in patients with COVID-19, with most based on artificial intelligence and machine learning from computed tomography images and several diagnostic predictors such as age, body temperature, clinical signs and symptoms, complications, epidemiological contact history, pneumonia signs, neutrophils, lymphocytes, and CRP levels (Li et al., 2020; Lopez-Rincon et al., 2020). Recently, by applying a proteomic and metabolomic measurement prediction model, COVID-19 patients that may become severe cases were identified (Shen et al., 2020). Although these models all report promising predictive performance with high C-indices (Wynants et al., 2020), they also carry a high risk of bias according to the PROBAST bias assessment tool (Moons et al., 2019). This is because most prediction models have not excluded patients with severe comorbidities and had a high risk of bias for the participant group or used non-representative controls, making the prediction results unreliable. Here, using strict inclusion and exclusion criteria, we minimized selection bias. According to the results of the prediction model, most mRNAs were highly correlated with asymptomatic patients (**Figures 4K, S10**). Multi-omics features such as *TBXA2R, ALOX15, IL1B, IFIT2, BCL2A1, LSP1*, glycyl-L-leucine and l-aspartate were highly expressed in the asymptomatic group, and thus may potentially yield crucial diagnostic biomarkers for identifying asymptomatic COVID-19 patients. In the critical illness group, beside CRP which has already been used to monitor the severity of COVID-19, some immune-related features, such as EEF1A1, FGL1, LRG1, CD99, COL1A1, cholinesterase(18:3), monoacylglyceride(18:1), Cannabidiolic acid and beta-asarone were found to be highly expressed in the critical group. Using these features, we could optimize existing approaches to improve the accuracy and sensitivity of detection based on nucleic acid testing and predict asymptomatic patient prognosis more accurately. With the assistance of this machine learning model, we could help identify individuals with a high risk of poor prognosis in advance, and prevent progression in time to minimize individual, medical and social costs.

## Study Limitations

The limitations of this research are as follows: (1) We did not enroll a healthy population as a control group, so conclusions made in this study are only limited to differences in COVID-19 severity. However, it is worth emphasizing that our research focused on the diversities and similarities in consecutively severe COVID-19. All stages of COVID-19 were included in our study design, in an attempt to identify key clues or biomarkers to distinguish disease severity and help prevent disease progression. (2) We excluded patients with comorbidities. As aforementioned, comorbidities including cancer history, hypertension, diabetes, cardiovascular disease, and respiratory diseases can affect COVID-19 progression. However, it remains unclear how these comorbidities affect COVID-19 progression and the relative weight of these comorbidities to progression. Thus, it would be risky to apply a specific prediction model to all COVID-19 patients. In conclusion, our study presented a panoramic landscape of blood samples within a large cohort of COVID-19 patients with various severities from asymptomatic to critically ill. Through trans-omics analyzing, we uncovered multiple novel insights, biomarkers and therapeutic targets relevant to COVID-19. Our data provided valuable clues for deciphering COVID-19, and the underlying mechanism warrant further pursuits.

## Data Availability

The data that support the findings of this study, including the genome-wide association test summary statistics, expression matrices for multi-omics have been deposited in CNSA (China National GeneBank Sequence Archive) in Shenzhen, China with accession number CNP0001126 (https://db.cngb.org/cnsa/).

## Acknowledgements

The study was supported by funding from National University Basic Scientific Research Special Foundation (2020kfyXGYJ00), China National GeneBank (CNGB) and Guangdong Provincial Key Laboratory of Genome Read and Write (No. 2017B030301011), Natural Science Foundation of Guangdong Province (2017A030306026), Funds for Distinguished Young Scholar of South China University of China (2017JQ017). We would like to thank Shangbo Xie, Yuying Zeng, Chengcheng Sun, Wendi Wu, Yan Li, Siyang Liu from BGI for helpful discussions of the results and advices. We would like to thank Ashley Chang for manuscript editing and data visualization.

## Author Contributions

D.M, X.J, G.C, C.S, L.W, P.W contributed to project conceptualization. D.M, X.J, X.X, S.L, J.W, H.Y contributed to the supervision. P.W, W.D, P.W, H.H, K.L, E.G, J.L, B.Y, J.F, L.H, Z.S, L.F, J.W, T.W, H.W, J.C, H.X, Y.M, Y.L contributed to sample collection. P.W, D.C, W.D, P.W, H.H, Y.B, Y.Z, K.L contributed to data analysis coordination. Y.R, Y.Z, K.H, W.S, Y.Z, H.L contributed to WGS, RNA-seq, LC-MS experiments. S.X, J.J, P.D, H.W, J.Q, F.W, J.Z, S.W, X.W, X.D, L.L, L.L, C.C, Z.Z contributed to RNA-seqanalysis M.H, Y.S contributed to miRNA-mRNA, lncRNA-mRNA interaction networks. Y.R, Y.Z, K.H, W.S, P.D, H.W, J.Q, F.W, J.Z, S.W, X.W, X.D, L.L, L.L, C.C contributed to proteomic analysis. Y.R, Y.Z, K.H, W.S, P.D, H.W, J.Q, F.W, J.Z, S.W, X.W, X.D, L.L, L.L, C.C metabolites analysis Y.Y, Y.R, Y.Z, K.H, W.S, P.D, H.W, J.Q,. W, J.Z, S.W, X.W, X.D, L.L, L.L, C.C contributed to lipids analysis Y.B contributed to machine learning model design and validation. Y.T, P.D, H.W, J.Q, F.W, J.Z, S.W, X.W, X.D, L.L, L.L, C.C contributed to data visualization Y.S, Y.Y, Z.Z, T.L, L.T, S.Z, L.Z, L.C, Y.W, X.M, F.C contributed to data interpretation. Y.Z contributed to data deposition. D.M, X.J, P.W, D.C, W.D, P.W, H.H, Y.B, Y.Z, K.L, L.W, C.S, G.C contributed to writing the original draft.

## Materials and Methods

### Ethics Statement

This study was reviewed and approved by the Institutional Review Board of Tongji Hospital, Tongji Medical College, Huazhong University of Science and Technology (TJ-IRB20200405). All the enrolled patients signed an informed consent form, and all the blood samples were collected using the rest of the standard diagnostic tests, with no burden to the patients.

### Patients Enrollment and Sample Preparation

Blood samples for 231 COVID-19 patients without any comorbidities were collected from Tongji Hospital and Union Hospital of Huazhong University of Science and Technology, Xiangyang Central Hospital, Hubei University of Arts and Science and Hubei Dazhong Hospital of Chinese Traditional Medicine from 19th February, 2020 to 26th April, 2020. Flowchart of patient selection for this study were shown in **Figure S1**. The demographic data and laboratory indicators were shown in **Table S1** and **S2**. The mean age of the patients was 46.7 years old (Standard Deviation=13.5), and the ratio of male to female was 1.12:1. All these patients were diagnosed following the guidelines for COVID-19 diagnosis and treatment (Trial Version 7) released by the National Health Commission of the People’s Republic of China. The patients were classified into four groups according to their disease severity: critical, severe, mild, and asymptomatic. The critical disease was defined as fulfilling at least one of the following conditions: (1) acute respiratory distress syndrome (ARDS) requiring mechanical ventilation, (2) shock, (3) combining with other organ failure requiring ICU admission. Severe disease met at least one of the following conditions: (1) respiratory rate ≥ 30 times/min, (2) oxygen saturation ≤93% at resting state, (3) arterial partial pressure of oxygen (PaO2)/fraction of inspired oxygen (FiO2) ≤300 mmHg, (4) pulmonary imaging examination showed that the lesions significantly progressed by more than 50% within 24-48 hours. Mild patients were defined as having fever, respiratory symptoms, lung imaging evidence of pneumonia. The patients with normal body temperature, without any respiratory symptoms were defined as asymptomatic. The definition of each severity was consistent with the previous article (Zhang et al., 2020). All Ethylenediaminetetraacetic acid disodium salt (EDTA-2Na)-anticoagulated venous blood samples were separated by centrifuge at 3,000 rpm, room temperature for 7min after standard diagnostic tests, the whole blood cells were stored at -80°C, 200 μL aliquot of serum were added 800μL ice-cold methanol, mixed well and stored at -80°C, another 200 μL aliquot of serum were added 800μL ice-cold isopropanol, mixed well and stored at -80°C.

### Nucleic Acid Extraction

A 200 μL aliquot of each thawed whole blood cells was used to extract DNA using QIAamp DNA Blood Mini Kit (51304, Qiagen), following the manufacturer’s instructions. Total RNA was extracted from another 200 μL aliquot of blood cells using QIAGEN miRNeasy Mini Kit (217004, Qiagen) according to the manufacturer’s protocol. All the extraction was performed under Level III protection in the biosafety III laboratory.

### Sequencing Library Construction and Data Generation

The whole genome data was generated through the following steps: 1) DNA was randomly fragmented by Covaris. The fragmented genomic DNA were selected by Magnetic beads to an average size of 200-400bp. 2) Fragments were end repaired and then 3’ adenylated. Adaptors were ligated to the ends of these 3’ adenylated fragments. 3) PCR and Circularization. 4) After library construction and sample quality control, whole genome sequencing was conducted on MGI2000 PE100 platform with 100bp paired end reads.

Transcriptome RNA data was generated through the following steps: 1) rRNA was removed by using RNase H method, 2) QAIseq FastSelect RNA Removal Kit was used to remove the Globin RNA, 3) The purified fragmented cDNA was combined with End Repair Mix, then add A-Tailing Mix, mix well by pipetting, incubation, 4) PCR amplification, 5) Library quality control and pooling cyclization, 6) The RNA library was sequenced by MGI2000 PE100 platform with 100bp paired-end reads.

Small RNA data was generated through the following steps: 1) Small RNA enrichment and purification, 2) Adaptor ligation and Unique molecular identifiers (UMI) labeled Primer addition, 3) RT-PCR, Library quantitation and pooling cyclization, 4) Library quality control, 5) Small RNAs were sequenced by BGI500 platform with 50bp single-end reads resulting in at least 20M reads for each sample.

### WGS Data Analysis and Joint Variant Calling

Whole genome sequencing data was processed using the Sentieon Genomics software (version: sentieon-genomics-201911) (Freed et al., 2017). Pipeline was built according to the best practice’s workflows for germline short variant discovery described in https://gatk.broadinstitute.org/. Sequencing reads were mapped to hg38 reference genome using BWA algorithm (Li and Durbin, 2009). After duplicates marking, InDel realignment and base quality score recalibration (BQSR), per-sample variants were called using the Haplotyper algorithm in the GVCF mode. Then the GVCFtyper algorithm was used to perform joint-calling and generate cohort VCF. Variant Quality Score Recalibration was performed using Genome Analysis Toolkit (GATK version 4.1.2) (Van der Auwera et al., 2013). The truth-sensitivity-filter-level were set as 99.0 for both the SNPs and the Indels. Finally, variants with PASS flag and quality score ≥ 100 were selected for further analysis.

### Genotype-Phenotype Association Analysis

PCA was performed using PLINK (v1.9) (Chang et al., 2015). Bi-allelic SNPs were selected based on the following criteria: minor allele frequency (MAF) ≥ 5%; genotyping rate ≥ 90%; LD prune (window = 50, step = 5 and r2 ≥ 0.5). A subset of 605,867 SNPs was used to perform PCA on the 203 unrelated individuals. We used rvtest (Zhan et al., 2016) to perform genotype-phenotype association analysis for 5,082,104 bi-allelic common SNPs with MAF > 5%. Gender, age and top 10 principal components were used as covariates for all the association tests. The qqman (Turner, 2014) and CMplot R packages (Yin, 2020) were applied to generate the Manhattan plot and quantile-quantile plot. We defined genome-wide significance for single variant association test as 5e^-8^, suggestive significance as 1e^-6^.

### QTL Analysis

We obtained matched proteomics, lipidomics, metabolomics, gene expression and SNP genotyping data for COVID-19 patients (n = 132). For the genotyping data, we removed outlier SNPs with MAF < 0.05. The QTL analysis (cis-eQTL analysis [local, distance < 10kb] for gene expression data, QTL analysis for proteomics, lipidomics, metabolomics data) was conducted using linear regression as implemented in MatrixEQTL (Shabalin, 2012). In this analysis, age and gender (1 for male and 2 for female) were considered as covariates. Associations with a p value less than 0.001 were kept, followed by FDR estimation using the Benjamini-Hochberg procedure as implemented in Matrix-QTL. QTL associations with an FDR-corrected p value < 5e^-8^ were considered significant (Frochaux et al., 2020).

### Gene Expression Analysis

RNA-seq raw sequencing reads were filtered by SOAPnuke (Li et al., 2008) to remove reads with sequencing adapter, with low-quality base ratio (base quality < 5) > 20%, and with unknown base (’N’ base) ratio > 5%. Reads aligned to rRNA by Bowtie2 (v2.2.5) (Langmead and Salzberg, 2012) were removed. Then, the clean reads were mapped to the reference genome using HISAT2 (Kim et al., 2015). Bowtie2 (v2.2.5) was applied to align the clean reads to the transcriptome. Then the gene expression level (FPKM) was determined by RSEM (Li and Dewey, 2011). Genes with FPKM > 0.1 in at least one sample were retained. Differential expression analysis was performed using DESeq2 (v1.4.5) with gender and age as confounders. Differential expressed genes were defined as those with Benjamini Hochberg adjusted p value < 0.05 and fold change > 2. GO enrichment analysis was performed using clusterProfiler (Yu et al., 2012). GO BP terms with an FDR adjusted p value threshold of 0.05 were considered as significant (Abdi, 2007).

Small RNA raw sequencing reads with low quality tags (which have more than four bases whose quality is less than ten, or have more than six bases with a quality less than thirteen.), the reads with poly A tags, and the tags without 3’ primer or tags shorter than 18nt were removed. After data filtering, the clean reads were mapped to the reference genome and other sRNA database including miRbase, siRNA, piRNA and snoRNA using Bowtie2 (Langmead and Salzberg, 2012). Particularly, cmsearch (Nawrocki and Eddy, 2013) was performed for Rfam mapping. The small RNA expression level was calculated by counting absolute numbers of molecules using unique molecular identifiers (UMI, 8-10nt). MiRNA with UMI count lager than 1 in at least one sample were considered as expressed. Differential expression analysis was performed using DESeq2 (v1.4.5) (Love et al., 2014) with gender and age as confounders to control for the additional variation and the detection cutoff was set as adjusted P < 0.05 and log2 of fold change ≥ 1.

### Construction of mRNA-miRNA and mRNA-lncRNA Network

To investigate the post-transcriptional regulation, spearman correlation coefficients of mRNA-miRNA (**Table S7**) and mRNA-lncRNA were calculated (**Table S8**). Correlation pairs with coefficients < −0.5 in mRNA-miRNA or < −0.6 in mRNA-lncRNA were retained. MultiMiR was used to confirm the top pairs of mRNA-miRNA by performing miRNA target prediction (Ru et al., 2014). The mRNA-miRNA and mRNA-lncRNA networks were visualized using Cytoscape (**Figure 2D**) (Shannon et al., 2003).

### Proteomics Analysis

The sera samples were inactivated at 56°C water bath for 30min and followed by processing with the Cleanert PEP 96-well plate (Agela, China). According to the manufacturer’s instructions, high-abundance proteins under a denaturing condition were removed (Lin et al., 2020). The Bradford protein assay kit (Bio-Rad, USA) was used to determine the final protein concentration. The proteins were extracted by the 8M urea and subsequently reduced by a final concentration of 10mM Dithiothreitol at 37°C water bath for 30min and alkylated to a final concentration of 55mM iodoacetamide at room temperature for 30min in the darkroom. The extracted proteins were digested by trypsin (Promega, USA) in 10 KD FASP filter (Sartorious, U.K.) with a protein-to-enzyme ratio of 50:1 and eluded with 70% acetonitrile (ACN), dried in the freeze dryer.

DIA (Data Independent Acquisition) strategy was performed by Q Exactive HF mass spectrometer (Thermo Scientific, San Jose, USA) coupled with an UltiMate 3000 UHPLC liquid chromatography (Thermo Scientific, San Jose, USA). The 1μg peptides mixed with iRT (Biognosys, Schlieren, Switzerland) were injected into the liquid chromatography (LC) and enriched and desalted in trap column. Then peptides were separated by self-packed analytical column (150μm internal diameter, 1.8μm particle size, 35cm column length) at the flowrate of 500 nL/min. The mobile phases consisted of (A) H2O/ACN (98/2,v/v) (0.1% formic acid); and (B) ACN/H2O (98/2,v/v) (0.1% formic acid) with 120 min elution gradient (min, %B): 0, 5; 5, 5; 45, 25; 50, 35; 52, 80; 55, 80; 55.5, 5; 65, 5. For HF settings, the ion source voltage was 1.9kV; MS1 range was 400-1250m/z at the resolution of 120,000 with the 50 ms max injection time(MIT). 400-1250 m/z was equally divided into 45 continuous windows MS2 scans at 30,000 resolution with the automatic MIT and automatic gain control (AGC) of 1E6. MS2 normalized collision energy was distributed to 22.5, 25, 27.5.

The raw data was analyzed by Spectronaut software (12.0.20491.14.21367) with the default settings against the self-built plasma spectral library which achieved deeper proteome quantification. The FDR cutoff for both peptide and protein level were set as 1%. Next, the R package MSstats (Choi et al., 2014) finished log2 transformation, normalization, and p-value calculation.

### Metabolomics Analysis

The 100μl sera of each sample were transferred into the 96-well plate and mixed with 10μl SPLASH LipidoMixTM Internal Standard (Avanti Polar Lipids, USA) and 10μl home-made Internal Standard mixture containing D3-L-Methionine (100 ppm, TRC, Canada), 13C9-Phenylalanine (100ppm, CIL, USA), D6-L-2-Aminobutyric Acid(100ppm, TRC, Canada), D4-L-Alanine (100ppm, TRC, Canada), 13C4-L-Threonine (100ppm, CIL, USA), D3-L-Aspartic Acid (100ppm, TRC, Canada), and 13C6-L-Arginine (100ppm, CIL, USA). The 300μl pre-chilled extraction buffer of methanol/ACN (67/33, v/v) was added to the plasma sample then vortexed for 1 min and incubated at -20°C for 2 hours. After centrifugation at 4000 RPM for 20 min, 300ul supernatants were taken and dried in the freeze dryer. The metabolites were dissolved in 150μl buffer of methanol/ACN (50/50, v/v) and centrifuged at 4000 RPM for 30min. Supernatants were injected into mass spectrometer.

Metabolomics data acquisition was completed using a same spectrometer, LC, and settings were set as lipidomics except for following parameters: the mobile phases of positive mode were (A) H2O (0.1% formic acid) and (B) methanol (0.1% formic acid). The mobile phases of negative mode were (A) H2O (10mM NH4HCO2) and (B) methanol /H2O (95/5, v/v) (10 mM NH4HCO2). Both positive and negative models used the same gradient (min, %B): 0, 2; 1, 2; 9, 98; 12, 98; 12.1, 2; 15, 2. The temperature of column was set at 45°C. MS1 range set as 70 -1050m/z. MS2 stepped normalized collision energy was distributed to 20, 40, 60.

The raw data was searched by Compound Discoverer 3.1 software (Thermo Fisher Scientific, USA) with different libraries including our self-built BGI library containing more than 3000 metabolites with corresponding detailed mass spectrum data. After quantification, subsequent processing steps were finished by metaX as same as lipidomics analysis.

### Lipidomics Analysis

The 100 μl sera of each sample was transferred into the 96-well plate and mixed with 10 μl SPLASH LipidoMixTM Internal Standard (Avanti Polar Lipids, USA). The 300μl pre-chilled Isopropanol (IPA) was added to the plasma sample and vortex for 1 min and incubated at -20°C overnight. Then samples were centrifuged at 4000 RPM for 20min while proteins precipitated. The supernatants were used for MS analysis.

Lipidomics analysis was performed using Q Exactive mass spectrometer (Thermo Scientific, San Jose, USA) coupled with Waters 2D UPLC (waters, USA). The CSH C18 column (1.7μm 2.1*100mm, Waters, USA) was used for separation with following elution gradient (min, %B) consisted of (A) ACN/H2O (60/40, v/v) (10 mM NH4HCO2 and 0.1% formic acid) and (B) IPA/ACN (90/10, v/v) (10 mM NH4HCO2 and 0.1% formic acid): 0, 40; 2, 43; 2.1, 50; 7, 54; 7.1, 70; 13, 99; 13.1, 40; 15, 40. The temperature of column was set as 55°C, the injection value was set as 5μL, and the flowrate was set as 0.35mL/min. For HF settings, the samples were scanned twice in both positive and negative modes. The positive spray voltage was set as 3.80 kV and negative spray voltage was set as 3.20 kV. MS1 range was 200-2000m/z at the resolution of 70,000 with the 100ms MIT and AGC of 3e6. The top3 precursors were set as trigger MS2 scans at the resolution of 17,500 with the 50ms MIT and AGC of 1E5. MS2 stepped normalized collision energy was distributed to 15, 30, 45. The sheath gas flow rate was set as 40 and the aux gas flow rate was set as 10.

The raw data was analyzed by Lipidsearch software Version 4.1 (Thermo Fisher Scientific, USA) which finished feature detection, identification and alignment. The following settings were applied: tolerance of mass shift, 5ppm; identification grade, A-D; filters, top rank; all isomer peak, FA priority, M-score, 5; c-score, 2.0; The export quantitative data from Lipidsearch was analyzed by R package metaX (Wen et al., 2017) which finished the normalization, correction of batch effect, and imputation of missing value.

For each patient in the cohort, we computed intensity for a given lipid complex class by summing up intensity of each lipid in the class. For each lipid complex class, the intensity value of each patient was further scaled by median value of intensity from mild patient group. We applied Mann-Whitney U-test (multiple comparisons correction with Bonferroni) to test statistically significant difference of scaled intensity of each lipid complex class between severity groups (**Figure S9**).

### Differential Expression of Proteins, Metabolites and Lipids

Expression data was first adjusted using robust linear model (RLM) for gender and age. The residuals following RLM were analyzed by Two-sided Mann-Whitney rank test for each pair of comparing group and p values were adjusted using Benjamini & Hochberg. Differentially expressed proteins, metabolites or lipids were defined using the criteria of adjust p value < 0.05 and fold change > 1.5.

### Clustering

Clustering was performed using the R package ‘Mfuzz’ after log2-transformation and Z-score scaling of the data. For mRNA from whole blood, genes differentially expressed in at least three out of the six comparison groups were clustered. For proteins, metabolites, lipids from sera, all the three analytes were clustered together.

### Pathway analysis

To annotate the proteins and metabolites in 7 clusters, gene ontology (GO) enrichment analysis were performed to obtain the enriched GO Biological Process terms of proteins in different clusters by clusterProfiler (Yu et al., 2012). And the 7 lists of metabolites in KEGG ID were classified into pathways by the Kyoto encyclopedia of genes and genomes (KEGG) database. The KEGG annotation was finished using in-house software.

### Correlation Network Analysis

Pairwise Spearman’s rank correlations were calculated using the r package ‘Hmisc’ and weighted, undirected networks were plotted with Cytoscape. Correlations with Bonferroni adjusted P values < 0.05 and absolute correlation coefficient >0.4 (**Figure S8**) were included and displayed via the Fruchterman-Reingold method. Nodes color indicate analytes type and their size represent the degree of the node.

### Data Preprocessing for Machine Learning

Extreme gradient boosting (XGBoost) (Chen and Guestrin, 2016), an ensemble algorithm of decision trees, was developed to predict patient severity status based on multi-omics data of mRNA transcripts (n=13323, mRNAs with FPKM >1 in at least one sample were retained), proteins (n=634), metabolites (n=814), lipids (n=742) from 135 patients (asymptomatic n=53, mild n=39, severe n=27, and critical n=16)using the open-sourced Python package (https://xgboost.readthedocs.io/en/latest/, version=1.0.0). We employed random stratified sampling to select 108 patients (80% of patient cohort) as the training set (asymptomatic n = 42, mild n = 31, severe n = 22, and critical n = 13), while the remaining 27 patients were used as the independent testing set (**Figure 4A)**. A fixed random number seed was used to ensure reproducibility of the results. The multi-omics data in training set was first normalized by centering and scaling for each sample to have mean zero and unit standard deviation. The estimated mean value and standard deviation for each feature from the training set were applied to the corresponding features in the testing phase afterwards.

## Feature Selection

Due to high dimensional multi-omics data and thus may decrease model’s performance if irrelevant features were included, we proposed a hybrid feature selection method to remove redundant and noise features. In this method, both mutual information (MI)-based technique and Boruta (Kursa and Rudnicki, 2010) algorithm were employed to obtain relevant subset of raw features. The MI-based technique was one of filter methods to select relevant features. It calculated weight by taking into account the relationship between features based on mutual information, and assigned the weight to each feature based on degree of relevance of features to class labels. We then selected 30% of features with the highest weights (Scikit-learn, version=0.23.1). The Boruta algorithm was one of wrapper methods to select subset of features based on a random forest machine learning algorithm that was used to measure feature importance. One feature was selected by Boruta only if its importance was greater than a threshold that was defined as the highest feature importance recorded among shadow features. The shadow features were obtained by permuting a copy of the real features across samples to destroy the relationship with the outcome. In Boruta, we applied random forest classifier with default parameters from Scikit-learn library except that class weight was specified due to imbalanced training set for each group of COVID-19 patent severity status. Python library BorutaPy (https://github.com/scikit-learn-contrib/boruta_py) was used to conduct Boruta algorithm using default parameters and a fixed random number seed. The final subset of relevant features was determined by computing intersection of subset features resulting from MI-based technique and Boruta algorithm. This procedure was repeated for each single-omics data and the final subset of relevant features was aggregated together for each sample.

### Model Training and Top Important Feature Identification

We performed a basic grid search algorithm with 5-fold cross validation to optimize XGBoost parameters while maximizing weighted F1 score because of the imbalanced training set (that is, the various number of samples in different patient group of COVID-19 severity). Consequently, the favorable values for the tuned XGBoost parameters were identified as follows: the maximum depth of trees was 8, number of decision trees was 55, minimum sum of instance weight needed in a child of a tree was 1, partitioning-leaf-node parameter was 0.4, subsample ratios of training instances for constructing each tree was 0.7, subsample ratios of columns was 0.9, learning rate was 0.05 and L1 regularization parameter was 0.005. We used softmax as learning objective function with predicted probability output per class due to the multi-class identification. The metrics of mean micro-average ROC curve with AUROC value and a mean micro-average PR curve with AUPR value were evaluated as the overall classifier performance when comparing one class to all others during the 5-fold cross validation for 100 iterations. In case of class imbalance, we calculated weight for each class and assigned each sample with corresponding class weight in the training set. After obtaining the favorable parameter values, the XGBoost model was trained using the entire training set.

We applied the SHAP (SHapley Additive exPlanations) (Lundberg and Lee, 2017; Lundberg et al., 2020) approach to measure feature importance for the XGBoost model. SHAP was a unified method to explain machine learning prediction based on game theoretically optimal Shapley values. To explain the prediction of a sample by the ML model, SHAP computed the contribution of each feature to the prediction, which was quantified using Shapley values from coalitional game theory. The Shapley value was represented as an additive feature attribution method, providing the average of the marginal contributions across all permutations of features and distribution of model prediction among features. As an alternative to permutation feature importance, SHAP feature importance was based on magnitude of feature attributions. The absolute Shapley values per feature across the data was further averaged as the global importance was needed. We ranked the features importance in descending order and picked the top 60 most important features. The stacked bar indicated the average impact of the feature on model output magnitude for different classes. We used the Python library to implement the SHAP algorithm (https://github.com/slundberg/shap). We re-trained the final XGBoost model based on the top 60 important features with the favorable model parameters using the entire training set.

### Machine Learning Model Evaluation

We evaluated the performance of the final XGBoost model as follows. We first normalized multi-omics data from the unseen 20% independent testing set using the mean value and standard deviation obtained during the training phase. Subsequently, features were screened based on the top 60 important features, followed by classification process using the final XGBoost model. The performance metrics included ROC curves with AUROC values, PR curves with AUPR values for each class, while micro-average ROC curves with AUROC values and micro-average PR curves with AUPR values for overall. In addition, confusion matrices (predicted label as the index of maximum value of the predicted probability vector) and UMAP plots (with parameters of the number of neighbors being 10, the minimum distance between points being 0.5 and the distance metric being Manhattan) were also generated for evaluating the performance.

To compare the performance of model based on multi-omics data to that based on single-omics data, we trained XGBoost model for single-omics data using the same training protocol as multi-omics data, except that we only empirically picked top 30 important features to train the final single-omics based XGBoost model. Moreover, we selected 20 proteins and 4 metabolites mentioned in Guo’s method (Shen et al., 2020), where 2 proteins and 1 metabolite were not found in our data set while 2 metabolites were greater than level 3 that were removed from our analysis. We trained XGBoost model using these 24 features with the same training protocol. Those models were evaluated on the unseen 20% independent testing set and calculated the same the performance matrices as mentioned above (**Figure S14**).

To further investigate the top 60 important features, we applied Mann-Whitney U-test (multiple comparisons correction with Bonferroni) to test statistically significant difference of each normalized features between severity groups (**Figure S10-S13**).

